# Growth kinetics of high-grade serous ovarian cancer using longitudinal clinical data - implications for early detection

**DOI:** 10.1101/2024.11.18.24317171

**Authors:** Bharath Narayanan, Thomas Buddenkote, Hayley Smith, Mitul Shah, Susan Freeman, David Hulse, Gabriel Funingana, Marie-Lyne Alcaraz, Mireia-Crispin Ortuzar, James Brenton, Paul Pharoah, Nora Pashayan

## Abstract

High-grade serous ovarian cancer (HGSOC) is the most lethal gynaecological cancer with patients routinely diagnosed at advanced stages with widespread disease. Evidence from screening trials indicates that early diagnosis may not reduce cancer-related deaths, possibly due to an underestimation of the true extent of the disease at screening. We aim to characterise the growth kinetics of HGSOC to understand why early detection has failed so far and under what conditions it might prove fruitful.

We analysed a dataset of 597 patients with a confirmed HGSOC diagnosis, and identified 37 cases with serial CT scans. We calculated the growth rates of lesions in the ovaries/pelvis and the omentum and estimated the time to metastasis using a population-level Gompertz model. Finally, we simulated ultrasound and CA125 based screening in a virtual population of patients.

Growing lesions in the ovaries and the omentum doubled in volume every 2.3 months and 2 months respectively. At both sites, smaller lesions grew faster than larger ones. The 12 cases with growing lesions in both disease sites had a median interval of 11.5 months between disease initiation and the onset of metastasis. Our simulations suggested that over 33% of patients would develop metastases before they could be screen detected. The remaining patients provided a median window of opportunity of only 4.7 months to detect the tumours before they metastasised.

Our results suggest that HGSOC lesions have short time to metastasis intervals, preventing effective early detection using current screening approaches.

## Introduction

Most patients with ovarian cancer are diagnosed at late stages with poor prognosis^1^; the 5-year survival rates of these individuals is less than half that of those diagnosed at early stages^2^. High-grade serous ovarian cancer (HGSOC) is the commonest histotype and is responsible for the majority of advanced-stage diagnoses and cancer-related deaths^3^. HGSOC originates in the fallopian tube as pre-cancerous serous tubal intraepithelial carcinoma (STIC) lesions^4^. They then detach from the fallopian tube and spread non-hematogenously to the ovaries, peritoneum, omentum and the lymph nodes^5^ where the disease can grow to a large volume without causing symptoms. Early detection initiatives aim to reduce mortality by detecting HGSOC as early as possible. However, screening for ovarian cancer is a challenging task, as evidenced by the lack of mortality benefit in recent trials. The United Kingdom Collaborative Trial of Ovarian Cancer Screening (UKCTOCS)^6^ found that a stage-shift in cancer diagnosis due to screening did not reduce cancer-related deaths over a median 16-year follow up period. Similarly, the Prostate, Lung, Colorectal and Ovarian cancer screening (PLCO) trial^7^ in the United States found no meaningful reduction in mortality through ultrasound (US) based screening. Consequently, both trials recommended against introducing screening programmes for ovarian cancer.

These findings suggest that by the time HGSOC can be screen-detected, it is already too late for most patients, regardless of the stage at which they are diagnosed. The success of screening programmes is linked to the growth rate of lesions and the interval between tumour initiation and metastatic dissemination. The lethality of HGSOC makes it challenging to gather sufficient longitudinal measurements to characterise its growth kinetics. Researchers have thus used mathematical models to infer the growth kinetics using the little data that is available. Brown and Palmer used the volumes and stage distributions of occult cancers identified during prophylactic bilateral salping-oopherectomy to estimate the tumour volume doubling time and the time spent by tumours in early stages before progression^8^. Danesh et al. used the growth rates from Brown and Palmer to calculate the interval between the earliest possible screen detection using US and the initiation of metastases using a branching process model^9^. Havrilesky et al. took a different approach and fitted a two-phenotype multi-state model to the stage distribution and outcomes from individual case data to identify the time spent by indolent (type-I) and aggressive (type-II) ovarian cancers in stage I/stage II^10^. Recent studies have been tailored to the growth kinetics of HGSOC in particular. Botesteanu et al. estimated the doubling time of HGSOC using volumes of diseased ovaries from 9 patients at single time points along with assumptions about the size of a normal ovary. They then developed a stochastic Gompertz model of HGSOC to identify the interval between a tumour being detectable through US and it reaching a life-threatening size^11^. Bedia et al. used longitudinal CA125 data from 504 women in the UKCTOCS trial to infer the growth rate of those HGSOC tumours that secreted CA125^12^. They also reported the preclinical detectable period (PCDP) between noticing a change in CA125 dynamics and eventual clinical diagnosis.

None of the aforementioned studies report the observed growth rates from two or more volume measurements. Although Bedia et al. used multiple biomarker measurements in their study, CA125 is a poor proxy for primary disease burden as we do not know the contribution from healthy tissue and each disease site. Additionally, Havrilesky et al. calculated the duration spent in early stages using staging/incidence data without information on the volume of metastatic disease. Therefore, these values are not true reflections of the time interval before metastasis as we do not know if the stage I/II cases in these datasets contained micro-metastases at diagnosis. In a similar vein, the PCDP reported by Bedia et al. does not tell us the window for detecting tumours before metastasis.

In this paper, we address the gaps in current knowledge by characterising the growth kinetics of lesions in the ovaries/pelvis and the omentum using longitudinal volumetric data from 37 HGSOC patients. The two disease sites were chosen as they account for most of the disease burden and are generally the most frequent locations for HGSOC^13^. Our work distinguishes itself from previous studies by reporting (i) the observed tumour growth rates between two volume measurements and (ii) the time interval between disease initiation and the onset of metastasis, using the growth kinetics of lesions in both disease sites.

## Methods

### Data

We used clinical data from a prospective cohort study, Cambridge Translational Cancer Research Ovarian Study 04^14^ (CTCR-OV04), designed to investigate the mechanisms of treatment response in ovarian cancer. We obtained access to individual data from 597 women in this cohort with histologically confirmed HGSOC, enrolled between September 2010 and September 2022. Sixty-six patients had multiple CT scans with contrast before treatment, making them suitable for modelling tumour growth kinetics. We included three more patients with no confirmed histotype resulting in a total of sixty-nine candidates for analysis. We only considered those cases with clinically observed primary disease and at least two scans taken with contrast at a 2mm slice thickness; forty-four cases satisfied these criteria.

### Patient and public involvement

We used pseudonymised and redacted data collected as part of the OV04 case-cohort study for this work. There was no patient or public involvement in the conceptualisation or execution of this study.

### Scan segmentation

We applied a deep-learning algorithm called *ovseg*^15^ on the CT scans to automatically segment lesions in the ovaries/pelvis and the omentum. *Ovseg* is a state-of-the-art segmentation framework that was designed and finetuned to segment lesions in HGSOC. It demonstrated performance comparable to a junior radiologist and achieved a Dice similarity coefficient of up to 71+-12 for ovarian/pelvic lesions and 61+-24 for omental lesions on external test data.

Although *ovseg* was trained on images acquired at a slice thickness of 5mm, we used scans acquired at intervals of 2mm as this provided a higher resolution and also expanded the number of suitable cases. Given this change in resolution, we revalidated the method by comparing the volumes obtained using the two slice thicknesses for 38 scans that had both resolutions available and a volume of at least 0.05cm^3^.

### Volume calculation

We calculated the total tumour burden in each disease site from the binary segmentation mask outputted by *ovseg* using:

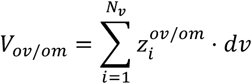

Where *N*_*v*_ is the total number of voxels in the scan sequence, *z*_*i*_ is 1 if the voxel is part of a lesion and 0 otherwise, and *dv* is the volume of a voxel.

### Doubling time calculation

We considered cases for doubling time calculations if they had at least 10% increase in volume between successive scans, and a minimum burden of 0.05 cm^3^ in either disease site; these requirements mitigated the chances of the results being skewed by artefacts.

We calculated the doubling time (TVDT) assuming an exponential growth model which provides the simplest way to capture the growth rate from two volume measurements.

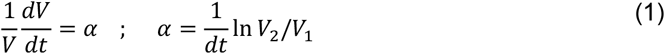

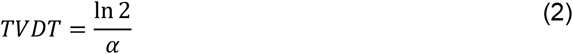

where *α* is the specific growth rate of the tumour, and *V*_2_ and *V*_1_ are the tumour volumes measured at a time interval of *dt*.

### Gompertz model of tumour growth

We sought a model that could best estimate the time of disease initiation from the available volumetric data. The constant growth rate assumption of the exponential model (equation 1) makes it unsuitable for this task as tumour growth has been shown to decrease with size until it becomes almost negligible at extremely large volumes^16–18^. Instead, we used a Gompertz model as it has been found to better mimic preclinical^18–20^ and clinical^21^ tumour growth patterns rate (equation 3).

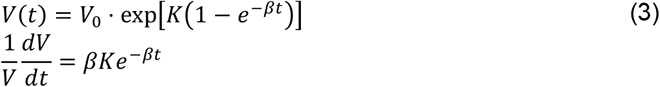

*V*_0_ is the initial volume, assumed to be a single cell (10^−9^ cm^3^), *β* is the decay constant, and 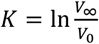, where *V*_*∞*_ is the carrying capacity of the tumour. We set *V*_*∞*_ as the maximal ovarian and omental volumes in our dataset, rounded to the nearest thousand.

The OV04 data consisted of two volume measurements, (*V*_1_, *V*_2_) along with the time interval, Δ*t* = *t*_2_ − *t*_1_, between the two scans. The time interval, *t*_1_, between disease initiation and the first scan is unknown and thus needed to be estimated along with *β* (equation 4).

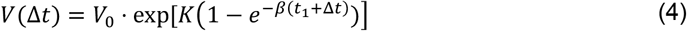

### Population modelling

We used a nonlinear mixed effects model (NLME) to fit the data and estimate the time between tumour initation and the first volume measurement in the two disease sites; this population approach to modelling enabled us to estimate the overall trend in growth kinetics while also accounting for inter-subject variability. NLME assumes that all individuals belong to a single population, and that their individual model parameters, ***θ***_***i***_, are distributed normally around the population average, ***θ***_***pop***_, with a variance matrix ***D***. The observation, 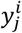, of individual *i* at time 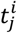; it is a function of ***θ***_***i***_ with an added error term, 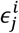, that follows a standard normal distribution and is scaled by *λ*.

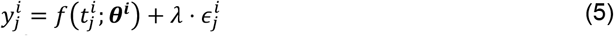

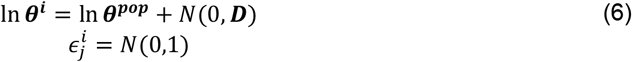

We used a reformulated version of the Gompertz equation with log-transformed parameters to ensure strictly positive values for *β* and *t*_1_ (equation 7).

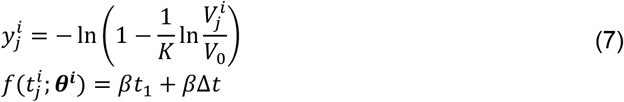

Here, ***θ*** = {*β, βt*_1_}, and the first observation at 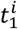 for each patient corresponds to 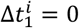. We estimated 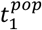 and the distribution of *t*_1_ from the fixed effects values and distributions of *β* and *βt*_1_. We compared the estimates of 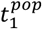 obtained using Gompertzian growth against those from an NLME with exponential growth (equation 1) where 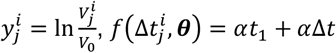,and ***θ*** = {*α, αt* }. However, we did not use these NLME estimates of exponential growth rates (*α*) while reporting the TVDT as we wanted to present the observed behaviour in the cohort of OV04 patients without extrapolating to a larger population.

In both the exponential and Gompertz growth cases, we first maximised the population likelihood using the stochastic approximation of expectation maximization (SAEM) algorithm^22^ to obtain an estimate of ***θ, D***, and *λ* and then fine-tuned the results by maximising the linearised log-likelihood using the quasi-Newton method^23^. Both algorithms were available as built-in functions in Matlab 2024^24^.

### Estimating the window-of-opportunity for screening in the OV04 dataset

We used the Gompertz model estimates of 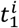 for each of the 12 cases with growing lesions in both disease sites to identify the WOO for screening. We first estimated the time to metastasis (Figure 2), 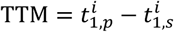, given by the interval between the initiation of disease in the primary (p) and secondary (s) disease sites. The primary site was not always the ovaries/pelvis as four cases were estimated to have disease initiated in the omentum first; we retrospectively found that these were cases of primary peritoneal cancer which is also classified as HGSOC^25^.

We then calculated the WOO for screening between the point at which the primary site reaches the US or CA125 detection thresholds of 0.5 cm^3^ and 0.015 cm^3^ respectively and the time of initiation of disease in the secondary site. These detection thresholds were informed by studies on the size distribution of image-detected tumours^26,27^ and a model-based estimate of the tumour burden at CA125 detection^28,29^.

### Estimating the window-of-opportunity for screening in an in-silico population

We simulated a virtual population of 10,000 patients to identify how many people could theoretically benefit from screening, and what the WOO would be in these individuals. For each case, we assigned the primary disease site with probabilities *p*_*om*_/*p*_*ov*_ and then sampled the Gompertz decay rate *β* from the corresponding distribution, while keeping 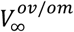 fixed. We used a value of 0.33 for *p*_om_ as 4 out of 12 cases in our cohort were expected to develop omental disease first; the results were agnostic to the choice of *p*_*om*_. Next, we sampled the size at which metastasis is seeded, *V*_*met*_, from the Gompertz estimate of the primary tumour at metastasis in the 12 patients with growing lesions in both sites. Finally, we calculated the time, *t*_*met*_, taken for the primary tumour to reach *V*_*met*_ and the WOO between the primary tumour reaching the CA125 / US detection limits and *t*_*met*_.

### Sensitivity analysis

Measurement error is inherent in segmentation and volume calculation. We studied the sensitivity of the estimated population mean, 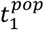, to these errors by applying 10% random Gaussian noise to the volumes before fitting the model. We repeated this 20 times and calculated the coefficient of variation (CV) of 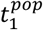 across these runs.

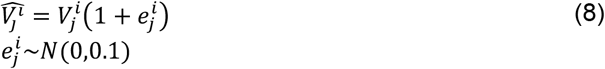

## Results

### Doubling times of lesions in the ovaries and omentum

We segmented lesions for 44 patients with a new diagnosis of HGSOC who had evaluable CT imaging before neoadjuvant chemotherapy or surgical treatment. We compared the volumes calculated at 2mm and 5mm slice thicknesses for 38 scans across 20 patients; we observed a median difference of 3.3% (IQR: 1.6 – 8.4) and 3.8% (1.2 – 10.7) for the ovarian and omental lesions respectively. We calculated the TVDT for 37 out of the 44 cases that had growing lesions in at least one of the two disease sites (Table 1) - 14 had growing ovarian lesions only, 11 had growing omental lesions only, and 12 had growing lesions in both disease sites (Supplemental material 1, figure S1).

**Table 1:** Characteristics of the 37 patients with a growing lesion in either the ovaries/pelvis or the omentum.

| N | 37 |
| --- | --- |
| Age | 69.1 ± 10.4 years |
| Stage |  |
| I | 1 |
| II | 1 |
| III | 27 |
| IV | 6 |
| NA | 2 |
| gBRCA status |  |
| Wildtype | 22 |
| NA | 13 |
| gBRCA2m | 2 |

The 23 omental lesions grew faster than the 26 omental/pelvic lesions (p-value 0.17) with a median TVDT of 2 months (IQR: [0.6, 2.6]) as compared to 2.3 months (IQR: [1.3, 4.2]) (Figure 1a). In both disease sites, lesions smaller than the median volume grew faster than those that were larger (Figure 1b); however, only the omental lesions displayed a statistically significant difference (p-value 0.026).

**Figure 1:**
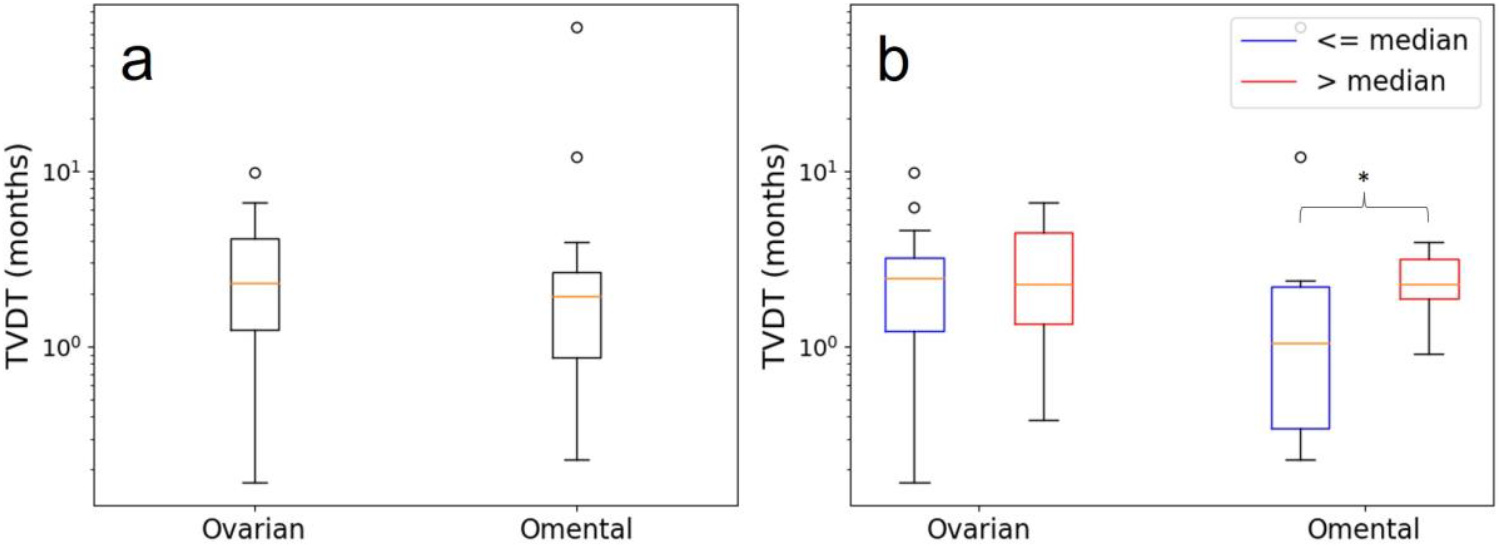
Volume doubling times (TVDT) a) by tumour site (26 ovary/pelvis and 23 omentum) and b) by tumour site and tumour volume at diagnosis - above (red) and below (blue) the median volume.

### Estimated time between disease initiation and diagnosis

The estimated population-level age of the tumour at first scan, 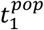,was 21 months for ovarian/pelvic lesions and 18 months for omental lesions. These values are substantially lower than those calculated using an exponential growth assumption – 84 months (IQR 42 – 153) for ovarian/pelvic lesions and 69 months (IQR 30 – 97) for omental lesions. This discrepancy is due to the exponential decay term in the Gompertz model; a tumour displaying a growth rate, *α*, between *V*_1_ and *V*_2_ will posess a higher growth rate at smaller volumes.

### Window-of-opportunity for detection using ultra-sound or CA125 based screening

We estimated a median TTM of 11.5 months (IQR 6.8 – 20) between the initiation of disease in the primary and secondary sites for the 12 cases with growing lesions in both disesae sites; this represents the most optimistic WOO for screening and suggests that half the cancers cannot be caught by annual screening even at a detection resolution of one cell. Moreover, the secondary disease is expected to have initiated before the primary reaches the CA125/US detection thresholds for 4 out of 12, and 6 out of 12 cases respectively (Figure 2b). Out of the eight cases that do reach the lower of the two thresholds before metastasis (Figure 2a), six had a WOO of less than 6.5 months.

**Figure 2:**
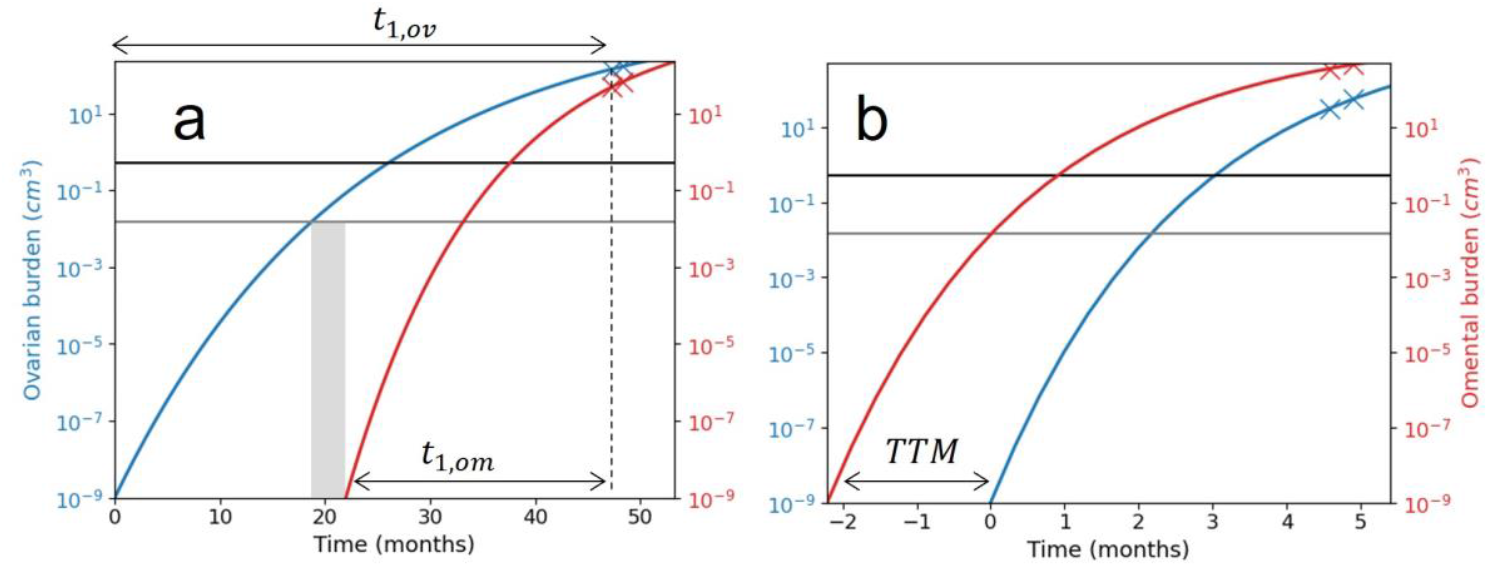
Gompertz growth profiles showing two cases: a) one that is expected to have developed an ovarian/pelvic lesion first and b) the other an omental lesion first. The grey and black horizontal lines denote the CA125 (0.015 cm3) and US detection limits (0.5cm3). In both cases, the secondary site is expected to have started growing before the primary lesion reaches the US detection threshold. However, CA125 based detection provides a theoretical WOO of around 3 months (grey rectangle) for the case with the ovarian primary (a).

Our simulations of a larger population yielded similar conclusions, with only 67.1% and 50% of the virtual patients expected to reach the CA125 and US detection limits before metastasis. The median WOO for these individuals was 4.7 months (IQR 2.2 – 10.3) and 2 months (IQR 0.5 – 4.5) using CA125 and US based screening respectively.

### Sensitivity of model estimates to errors in measurement and choice of maximal disease burden

The estimated fixed effects parameter, 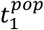, had a coefficient of variation (CV) of 14.8% and 8.5% for ovarian and omental lesions respectively when subjected to noisy measurements. However, the distributions themselves were visibly different (figure 3a and 3b), suggesting that estimates of the population mean are robust to measurement errors even when individual estimates can be different.

**Figure 3:**
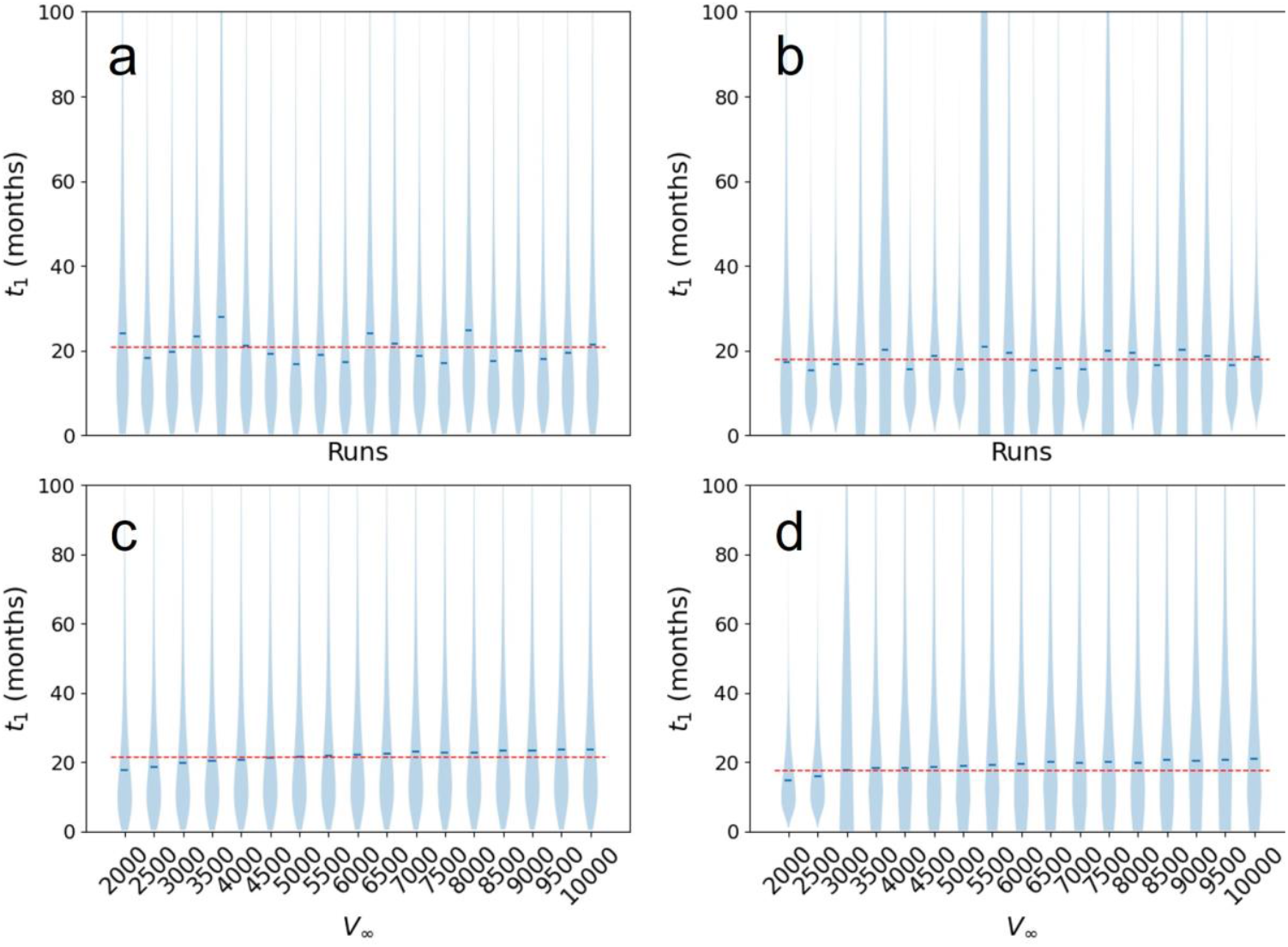
Sensitivity of the t1 estimates to measurement errors and the choice of V_∞_. The violin plots in the top row show the log-normal distributions of t_1_ estimates for each of the 20 runs with 10% Gaussian noise. We observed a CV of 14.8% and 8.5% in the estimated population-level 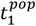 (blue horizontal lines) for (a) ovarian and (b) omental lesions respectively. The red dashed lines represent the estimated 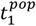 without any noise. The bottom row shows the moderate rise in the estimated t_1_ as the assigned V_∞_ for (c) ovarian and (d) omental lesions is increased from 2000 cm3 to 10,000 cm3. The CV in 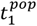 was 8.3% and 8.7% for ovarian and omental sites respectively

In a similar vein, the estimated 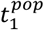 had a CV of 8.3% and 8.7% when the *V*_∞_ was varied between 2000 and 10000 cm^3^ in the ovarian and omental sites respectively. This variation dropped to 3.2% and 4.8% for carrying capacities at or above the values set for the ovarian (5000cm^3^) and omental (3000cm^3^) lesions respectively, indicating a negligible impact of the choice of *V*_∞_ on our findings.

## Discussion

Cancer growth kinetics dictate the probable success of early detection and clinical interventions. In this study, we used longitudinal clinical data from 37 HGSOC patients to understand how fast aggressive ovarian cancer tumours grow in the ovaries/pelvis and the omentum, how soon after tumour initiation they spread, and what the likely interval is for screening. We found that lesions in the ovaries/pelvis and the omentum doubled in volume every 2.3 months and 2 months respectively while the median TTM in the cohort was 11.5 months. Simulations of a population of tumours suggest that only 67% of tumours can be caught before metastasis using CA125 based screening with a median WOO of 4.7 months for early detection in these cases. US based screening is predicted to carry even lower potential for early detection with only 50% of the cases reaching the detection limit before metastasis, with a median WOO of 2 months. These findings not only corroborate evidence of the inefficacy of annual screening, they also suggest that no screening programme is likely to catch cancers before metastasis, given current thresholds of detection.

Our reported values for the kinetic parameters are consistent with those obtained using other approaches. The reported median TVDT for ovarian lesions falls within the ranges reported by Brown and Palmer^8^ (2.5 months), Botesteneau et al.^11^ (1.73 months), and Bedia et al.^12^ (2.9 months). Similarly, the TTM between primary and secondary disease initiation compares well with the period of 12.9 months^10^ spent in stage I/II as estimated by a natural history model of aggressive (type-II) cancers. We find it encouraging that studies based on different types of data and different models arrive at similar estimates of the growth kinetics of HGSOC. However, our values for the TTM and WOO cannot be directly compared to the preclinical detectable periods (PCDP) of 11.9 months reported by Bedia et al., 2 years reported by Ishizawa et al.^30^ or 2.9 years reported by Botesteanu et al^11^. These values represent the interval between the earliest possible screen detection and clinical diagnosis. In contrast, our values represent the time between disease initiation (TTM) or screen detection (WOO) and the *onset* of metastasis which could occur earlier than the presentation of clinical symptoms.

We used two different modelling approaches for this work; the exponential model fitted to individual patient data enabled us to report the observed TVDT for the OV04 patient cohort without any extrapolation while the population-level Gompertz model was better equipped for backprojecting the course of disease and estimating the likely WOO. Our sensitivity analyses provide confidence that the reported population-level WOO will qualitatively hold under measurement errors and different choices of carrying capacity. The analyses also show that population-level Gompertz growth parameters are robust to observation errors even if individual estimates are susceptible to such perturbations.

The results of our work have important caveats. We use a small sample size of 37 patients from a single site and empirical growth models for estimating the growth kinetics of HGSOC. We urge caution when extrapolating these findings to a more heterogeneous and global context despite the congruency of our estimated parameters with work done on a larger cohort^12^. We obtained the time since disease initiation by back-projecting a Gompertz model that has previously been shown to capture tumour growth in preclinical and clinical settings. Nevertheless, there is no clinical evidence that HGSOC in particular follows a Gompertzian growth in-vivo. Moreover, our reliance on just two measurements per individual means that any model can fit the data; there is no way to evaluate different models by validating the back projected volumes against additional measurements done in preclinical studies^19,31^. The limitation of having insufficient data is driven by a contradiction inherent to fast growing cancers – the speed of growth and dissemination that underpins their lethality also renders them difficult to observe and characterise. Numerical experiments suggest that faster growing tumours need fewer measurements to characterise their growth. However, we would need at least 3-4 measurements to accurately capture individual Gompertzian kinetics in the presence of even 5% measurement error^32^.

The above limitations notwithstanding, this is the first study that uses (i) longitudinal volumetric data to infer the growth rates of HGSOC in different sites, and (ii) growth models of both primary and metastatic disease to estimate the WOO for screening. Notably, the kinetic parameters and study conclusion that we obtained through volume-based mechanistic modelling are consistent with those using a natural history model and biomarker based kinetics. Our work provides further evidence of the challenges involved in ovarian cancer screening; it is likely that HGSOC grows too fast and spreads too soon for any meaningful screening-led reduction in mortality.

## Data Availability

All data produced in the present work are contained in the manuscript. Code for the simulations and model fitting are available upon reasonable request.

## Ethics Statement

This work uses anonymised patient data collected as part of the CTCR-OV04 study. The OV04 study was given ethical approval by the Institutional Ethics Committee (REC reference number 08 /H0306/61). Patients provided written consent to their anonymised data being used in secondary studies, such as this work, and for their anonymised data to be shared with other researchers. No other data or patient information was collected in this study.

## Data Access

The codes and the fitted models used in this study can be shared upon reasonable request.

## References

1 NHS England. Case-mix adjusted percentage of cancers diagnosed at stages 1 and 2 in England. 2022.

2 NHS England. Cancer Survival in England, cancers diagnosed 2016 to 2020, followed up to 2021. 2023.

3 Gaitskell K, Hermon C, Barnes I, Pirie K, Floud S, Green J et al. Ovarian cancer survival by stage, histotype, and pre-diagnostic lifestyle factors, in the prospective UK Million Women Study. Cancer Epidemiol 2022; 76. doi:10.1016/j.canep.2021.102074.

4 Labidi-Galy SI, Papp E, Hallberg D, Niknafs N, Adleff V, Noe M et al. High grade serous ovarian carcinomas originate in the fallopian tube. Nat Commun 2017; 8. doi:10.1038/s41467-017-00962-1.

5 Lisio MA, Fu L, Goyeneche A, Gao ZH, Telleria C. High-grade serous ovarian cancer: Basic sciences, clinical and therapeutic standpoints. Int J Mol Sci. 2019; 20. doi:10.3390/ijms20040952.

6 Menon U, Gentry-Maharaj A, Burnell M, Ryan A, Kalsi JK, Singh N et al. Mortality impact, risks, and benefits of general population screening for ovarian cancer: the UKCTOCS randomised controlled trial. Health Technol Assess (Rockv) 2023; : 1–81.

7 Pinsky PF, Yu K, Kramer BS, Black A, Buys SS, Partridge E et al. Extended mortality results for ovarian cancer screening in the PLCO trial with median 15 years follow-up. Gynecol Oncol 2016; 143: 270–275.

8 Brown PO, Palmer C. The preclinical natural history of serous ovarian cancer: Defining the target for early detection. PLoS Med 2009; 6. doi:10.1371/journal.pmed.1000114.

9 Danesh K, Durrett R, Havrilesky LJ, Myers E. A branching process model of ovarian cancer. J Theor Biol 2012; 314: 10–15.

10 Havrilesky LJ, Sanders GD, Kulasingam S, Chino JP, Berchuck A, Marks JR et al. Development of an ovarian cancer screening decision model that incorporates disease heterogeneity. Cancer 2011; 117: 545–553.

11 Botesteanu DA, Lee JM, Levy D. Modeling the dynamics of high-grade serous ovarian cancer progression for transvaginal ultrasound-based screening and early detection. PLoS One 2016; 11. doi:10.1371/journal.pone.0156661.

12 Bedia JS, Ian Jacobs HJ, Ryan A, Gentry-Maharaj A, Burnell M, Manchanda R et al. Estimating the Ovarian Cancer CA-125 Preclinical Detectable Phase, In-Vivo Tumour Doubling Time, and Window for Detection in Early Stage: An Exploratory Analysis of UKCTOCS. Preprint 2024.https://ssrn.com/abstract=4834024.

13 Crispin-Ortuzar M, Woitek R, Reinius MAV, Moore E, Beer L, Bura V et al. Integrated radiogenomics models predict response to neoadjuvant chemotherapy in high grade serous ovarian cancer. Nat Commun 2023; 14. doi:10.1038/s41467-023-41820-7.

14 Parkinson CA, Gale D, Piskorz AM, Biggs H, Hodgkin C, Addley H et al. Exploratory Analysis of TP53 Mutations in Circulating Tumour DNA as Biomarkers of Treatment Response for Patients with Relapsed High-Grade Serous Ovarian Carcinoma: A Retrospective Study. PLoS Med 2016; 13. doi:10.1371/journal.pmed.1002198.

15 Buddenkotte T, Rundo L, Woitek R, Escudero Sanchez L, Beer L, Crispin-Ortuzar M et al. Deep learning-based Segmentation of Multi-site Disease in Ovarian Cancer. Eur Radiol Exp 2023; 7. doi:10.1101/2023.01.10.22279679.

16 Steel GG, Lamerton LF. The Growth Rate of Human Tumours. Br J Cancer 1966; 20.

17 Spratt JA, von Fournier D, Spratt JS, Weber EE. Decelerating growth and human breast cancer. Cancer 1993; 71: 2013–2019.

18 Laird AK. Dynamics of Tumor Growth. Br J Cancer 1964; 18.

19 Benzekry S, Lamont C, Beheshti A, Tracz A, Ebos JML, Hlatky L et al. Classical Mathematical Models for Description and Prediction of Experimental Tumor Growth. PLoS Comput Biol 2014; 10. doi:10.1371/journal.pcbi.1003800.

20 Winsor CP. The Gompertz curve as a growth curve. Proceedings of The National Academy of Sciences 1932; 18.

21 Norton L. A Gompertzian Model of Human Breast Cancer Growth. Cancer Res 1988; 48: 7067–7071.

22 Comets E, Lavenu A, Lavielle M. Parameter estimation in nonlinear mixed effect models using saemix, an R implementation of the SAEM algorithm. J Stat Softw 2017; 80. doi:10.18637/jss.v080.i03.

23 Kuhn E, Lavielle M. Maximum likelihood estimation in nonlinear mixed effects models. Comput Stat Data Anal 2005; 49: 1020–1038.

24 The MathWorks Inc. MATLAB version: 24.1.0 (R2024a) Update 3. 2024.

25 Saida T, Tanaka YO, Matsumoto K, Satoh T, Yoshikawa H, Minami M. Revised FIGO staging system for cancer of the ovary, fallopian tube, and peritoneum: important implications for radiologists. Jpn J Radiol. 2016; 34: 117–124.

26 Prakash P, Cronin CG, Blake MA. Role of PET/CT in ovarian cancer. American Journal of Roentgenology. 2010; 194. doi:10.2214/AJR.09.3843.

27 Healy JC. Detection of peritoneal metastases. Cancer Imaging 2001; 1. doi:10.1102/1470-7330.2001.002.

28 Mathieu KB, Bedi DG, Thrower SL, Qayyum A, Bast RC. Screening for ovarian cancer: imaging challenges and opportunities for improvement. Ultrasound in Obstetrics and Gynecology. 2018; 51: 293–303.

29 Lutz AM, Willmann JK, Cochran F V., Ray P, Gambhir SS. Cancer screening: A mathematical model relating secreted blood biomarker levels to tumor sizes. PLoS Med 2008; 5: 1287–1297.

30 Ishizawa S, Niu J, Tammemagi MC, Irajizad E, Shen Y, Lu KH et al. Estimating sojourn time and sensitivity of screening for ovarian cancer using a Bayesian framework. JNCI: Journal of the National Cancer Institute 2024. doi:10.1093/jnci/djae145.

31 Vaghi C, Rodallec A, Fanciullino R, Ciccolini J, Mochel JP, Mastri M et al. Population modeling of tumor growth curves and the reduced Gompertz model improve prediction of the age of experimental tumors. PLoS Comput Biol 2020; 16. doi:10.1371/journal.pcbi.1007178.

32 Harshe I, Enderling H, Brady-Nicholls R. Predicting Patient-Specific Tumor Dynamics: How Many Measurements Are Necessary? Cancers (Basel) 2023; 15. doi:10.3390/cancers15051368.

