## Supplemental material for "Growth kinetics of high-grade serous ovarian cancer using longitudinal clinical data - implications for early detection"

### Note 1 – deriving the CI of $t_{1}$

Letting $q=\beta t_{1}$, we first calculate the population level $t_{1}^{pop}$

$$\left( \ln t_{1} \right)^{pop}=\left( \ln q \right)^{pop}-\left( \ln\beta\right)^{pop}$$

$$t_{1}^{pop}=\exp\left( \ln t_{1} \right)^{pop}$$

For the confidence interval (CI) of $t_{1}^{pop}$ we start by providing the formula for the standard error of the difference of two independent, normally distributed variables:

$$SE_{X-Y}=\sqrt{SE_{X}^{2}+SE_{Y}^{2}}$$

Here, $X-Y=\left( \ln t_{1} \right)^{pop}$, $X=\left( \ln q \right)^{pop}$ and $Y=\left( \ln\beta\right)^{pop}$

$$SE_{\left( \ln t_{1} \right)^{pop}}=\sqrt{SE_{\left( \ln q \right)^{pop}}^{2}+SE_{\left( \ln\beta\right)^{pop}}^{2}}$$

Now, the confidence interval of $\left( \ln t_{1} \right)^{pop}$ is

$$CI_{\left( \ln t_{1} \right)^{pop}}=\left[ \left( \ln t_{1} \right)^{pop}\mp t_{.975}\cdot SE_{\left( \ln t_{1} \right)^{pop}} \right]$$

We use the t-distribution as we only have 26 individuals for the ovarian lesions ($t_{.975}=2.06$) and 22 individuals for the omental lesions ($t_{.975}=2.074)$.

Finally, we estimate the CI for the population level estimate of $t_{1}$:

$$CI_{t_{1}^{pop}}=\exp CI_{\left( \ln t_{1} \right)^{pop}}$$

Note that we seek the CI estimate for $\exp\left( \ln t_{1} \right)^{pop}$and not the CI for the mean of $t_{1}$, $\mu_{t_{1}}=\exp(\mu_{\ln t_{1}}+\sigma_{\ln t_{1}}^{2})$. This is why we use the naïve approach to CI estimation here.

### Figure S1

*Ovarian* ***(a)*** *and omental* ***(b)*** *volumes from the 37 cases used in this study. The blue lines represent the 12 cases with growing lesions in both disease sites, while the black lines denote the cases with growing ovarian (14) or omental (11) lesions only. Cases coloured red were those that were discarded for volume calculation due to a lack of growth (<10%), or their extremely small size (<0.05cm^3^). Both axes are in log-scale to better distinguish the different magnitudes of volumes. One of the cases had a measured ovarian volume of 0 at the first scan (a-red, almost vertical line); we assigned a volume of 1e-16 as the first measurement so that this case could be plotted on the log-scale. We also assigned the first scan for all cases to be at 1 month at the first scan because 0 months cannot be plotted on the log scale.*

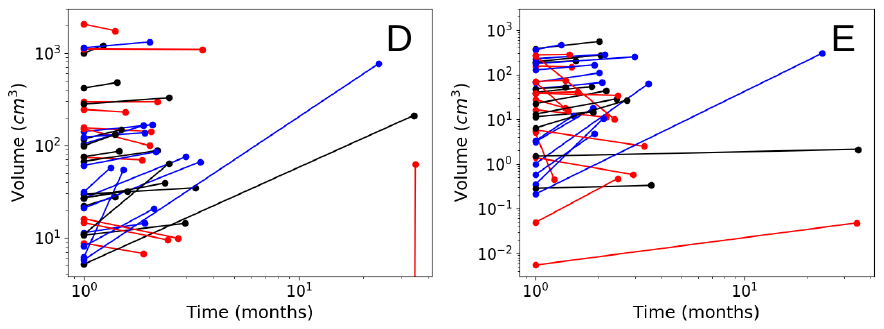
